# Rapid genome surveillance of SARS-CoV-2 and study of risk factors using shipping container laboratories and portable DNA sequencing technology

**DOI:** 10.1101/2022.02.25.22271277

**Authors:** Sara Farahi Bilooei, Dejana Jovicevic, Arash Iranzadeh, Anthony Thomas, Ivan Muscat, Cynthia Mpofu, Helene Steiner, Thomas Meany

## Abstract

In this paper we report on genome sequencing of 154 SARS-CoV-2 samples between June and July 2021 (Summer outbreak) in the Bailiwick of Jersey, a UK channel island. We have analysed extensive data collected on 598,155 RT-qPCR tests that identified 8,950 positive cases as part of public health surveillance from September 2020 to August 2021. Our study implemented an amplicon-based sequencing approach using the Oxford Nanopore Technology (ONT) portable device. This revealed the emergence of twelve AY sublineages and were clustered into the Delta sub-clades 21I and 21J. This was integrated alongside an existing RT-qPCR diagnostic laboratory to provide a sample-to-sequence turnaround time of approximately 30 hours with significant scope for optimisation. Owing to the geographic remoteness of the island from large scale sequencing infrastructure, this presents an opportunity to provide policy makers with near real-time sequencing findings. Our analysis suggests that age and sex remained a substantial risk factor for mortality. We observe viral loads are higher in advanced ages and unvaccinated individuals. The median age of SARS-CoV-2 positive individuals was higher during winter than the summer outbreak, and the contact tracing program showed that younger individuals stayed positive for longer.

## Introduction

The severe acute respiratory syndrome coronavirus-2 (SARS-CoV-2) first emerged in Wuhan, China and caused coronavirus disease of 2019 (COVID-19) ^1,2^. SARS-CoV-2 has spread with the evolution of its viral genome since its emergence in late 2019. The first SARS-CoV-2 genome sequence confirmed the virus as part of the betacoronavirus genus, belonging to the *Coronaviridae* family, consisting of single-strand positive-sense RNA ^1,3^. Since the publication of the first sequence at the beginning of 2020, viral genome sequencing has become a powerful tool to study the SARS-CoV-2 genome ^1,4^. SARS-CoV-2 genome variation study enables a comprehensive understanding of virus transmission, rate of mutations, track evolution, development of vaccines, and treatment ^5^. This virus constantly changes by distinguishable mutations that have emerged in different geographical locations, each promptly determined as the dominant variant within a few months of initial detection ^6,7^. The Alpha (B.1.1.7) variant was first identified in the UK in November 2020, and the Delta (B.1.617.2) variant was detected in India during October 2020 and quickly became a variant of concern (VOC)^8^. SARS-CoV-2 has a genome of 29,903 nucleotides, with 5’-cap structure and 3’ poly(A) tail ^9,10^. The 5’ terminus genome encodes nonstructural proteins (nsps) that are involved in the process of the virus infection cycle. The 3’ terminus encodes structural proteins (spike (S), envelope (E), membrane (M) and the nucleocapsid (N)) and several accessory proteins ^9,11^. The spike protein has a strong binding affinity to the human angiotensin-converting enzyme receptors (ACE2) and causes infection resulting in COVID-19 disease ^12,13^. The spike protein is highly immunogenic and acts as an essential target of vaccine development ^14,15^ and plays a crucial role in facilitating human to human transmission, even during the incubation period (asymptomatic stage) ^16,17^. The characteristic S protein mutations in the Delta variant accumulated five mutations in the N-terminal domain (NTD) (T19R, G142D, E156-, F157- and R158G), (L452R and T478K) in the receptor binding domain (RBD) region, (P681R) located in the furin-cleavage site ^18^, and (D950N) in the S2 region ^19,20^. Researchers have identified mutations and modifications associated with infectivity and reactivity to neutralise antibodies ^21^. A substitution in the spike (D614G), caused by a missense mutation of aspartate to glycine, is shared with the four significant VOCs: Alpha, Beta, Gamma and Delta strains. S: D614G emerged during the pandemic’s begining and quickly became common in Europe and North America ^22–24^. The first travel-associated positive case detected in the Bailiwick of Jersey was the 10th of March 2020. Being a small island with a population of 107,800, Jersey quickly established a free testing programme that ranked high amongst the most effective in Europe early in the COVID-19 pandemic. In September 2020, an OpenCell COVID-19 rapid testing laboratory was deployed inside a shipping container designed to diagnose the virus effectively. The laboratory carried out a reverse transcription-polymerase chain reaction (RT-qPCR) diagnostic workflow assay, which formed part of the Government of Jersey testing programme ^25^. Patient registration, swabbing, and diagnostic testing of COVID-19 (CONTAIN Jersey) had an average turnaround of 12 hours, and genome sequencing of SARS-CoV-2 took 18 hours by the MinION platform. The Oxford Nanopore MinION is a field-deployable, portable device capable of generating real-time, long-read sequences, facilitating the fastidious sequencing of viral genomes ^26,27^. The MinION generates high read coverage, cost-effective, and rapid sequencing. The cost of sequencing using the ONT device (MK1C) is presented in Supplementary Fig S1. This study performed sequencing of 154 SARS-CoV-2 positive samples to identify amino acid change(s) and the variants circulating in the community. We aimed to identify risk factors associated with an increased risk of mortality.

## Methods

### Data collection and source

The Government of Jersey (GoJ) performed primary data collection as public health surveillance from September 2020 to August 2021. The testing programme involved screening all incoming passengers at the ports of entry (air or boat) and any domiciled residents within the community requesting a SARS-CoV-2 test (via contact tracing or symptomatic reasons). All passengers aged 11 and over were asked to complete a safer travel registration form upon arrival, including relevant information such as COVID-19 vaccination record, symptoms (if any) and previous travel history. All passengers and everyone tested through the community were asked to identify all symptoms before testing. If the infected subjects presented no symptoms, these cases were classed as asymptomatic. In addition, qualitative sequencing data was received from GoJ with sequencing performed by Micropathology laboratory Ltd, where an independent secondary swab of positive individuals was sequenced. The data received from these genome sequences was in the form of notes providing several mutations and the variant of the samples.

### Statistical analysis

A dataset was created, including the age (aggregated into 10-year groupings), cycle threshold (Ct) values, travel history, reasons to seek test, and symptomatic infected cases. The data also included symptoms, described at the time of test, associated with SARS-CoV-2. Data were presented as mean, median [interquartile range (IQR)], or percentage as appropriate. The categorical variables were presented as numbers (percentages). The chi-square (χ2) test was used to compare the differences between the two groups. Kruskal-Wallace test was used to compare Ct values in age categories. The analysis was conducted based on calendar weeks or months. Spearman rank correlation was used to determine the correlation between viral load (Ct value used as a proxy) and age of direct contact positive cases.

### SARS-CoV-2 specimens

Positive SARS-CoV-2 specimens were extracted from 154 positive samples and tested at the Jersey OpenCell COVID-19 laboratory. Samples were oral-nasopharyngeal swabs collected from individuals in Jersey during June and July 2021 for genome sequencing.

### RNA isolation and RT-qPCR testing

The RNA was extracted using the MagMAX™ Express-96 Deep Well Magnetic Particle Processor (Thermo Scientific™ KingFisher™ Flex Robot, Thermo Fisher Scientific) using automation-ready protocols according to the supplier’s instructions. Detection of SARS-CoV-2 virus was performed using 2019-nCoV RealStar SARS-CoV-2 RT-qPCR Kit (Altona Diagnostics) by RT-qPCR assay (Quantstudio™ 2, Applied Biosystems) targeting B-betacoronavirus (B-βCoV) (E gene target) and SARS-CoV-2 specific RNA (S gene target). A positive result required at least one of the two-targeted regions to amplify at any Cts. A low Ct value determines a high level of detectable viral genetic material and vice versa.

### Whole Genome Sequencing

SARS-CoV-2-positive RNA extract of 154 samples was prepared and sequenced by the protocol developed by the ARTIC network ^28,29^. Briefly, the cDNA synthesis was performed on extracted RNA of SARS-CoV-2 positive samples with Ct number less than ≥33, using LunaScript RT SuperMix Kit (New England Biolabs). SARS-CoV-2 whole-genome amplification was performed by multiplex PCR using the ARTIC V3 primer scheme to generate 400-bp amplicons. The PCR products were diluted 5-fold and were end-prepped by Ultra II end-prep enzyme mix (NEB). Native barcode ligation was prepared from PCR barcoding Expansion 1-96 kit (EXP-PBC196, ONT). The amplicons were pooled and purified by AMPureXP (Beckman Coulter Life Sciences). The adaptor ligation was performed with NEBNext Quick T4 Ligase and AMII (ONT) and bead-purified. The DNA concentration was measured using the Qubit 2.0 instrument (Thermo Fisher). The library was loaded on a prepared R9.4 flow cell, multiplexing up to 48 samples per run.

### Basecalling, assembly, and variant calling

Basecalling was performed on fast5 raw sequence data generated with MinION MK1C using Guppy v.4.2.2 (ONT) with the high-accuracy base-call setting (model dna_r9.4.1_450bps_hac). The obtained data were demultiplexed using guppy_barcoder (v4.2.2) with barcodes separating into individual folders in fastq reads. Nanopore fastq files were assembled and aligned to the Wuhan-Hu-1 reference genome (NC_045512.2) using the Coronavirus Typing Tool on Genome Detective 1.136 https://www.genomedetective.com, a web-based software application. Genomes were submitted to the Nextclade web page https://clades.nextstrain.org, and those that did not pass the standard quality control parameters were filtered out. Frameshift mutations and misplaced stop codons were polished manually by aligning genomes to the reference using MAFFT ^30^ and visualising the alignment files by Aliview ^31^. Viral lineages were classified using the Pangolin (v3.1.16) ^32^ software tool http://pangolin.cog-uk.io.

### Sequencing laboratory in a shipping container

Previous work by Walker et al. (2020) presented an automated, UKAS accredited (Testing Laboratory No. 22071) COVID-19 diagnostic laboratory housed within a shipping container^25^. The nanopore sequencing was conducted in an adjacent laboratory also fitted in a shipping container. The sequencing laboratory requires a linear physical layout matching the workflow. The space of the container is divided into three different sections. Station A: contains two cabinets to avoid contamination. The RNA extracted from positive samples of SARS-CoV-2 are reverse transcribed. The cDNA mastermix should be added to the PCR plate in cabinet A and Viral RNA from samples must be added to the plate in cabinet B. The multiplex PCR Q5 HOTstart DNA Polymerase mastermix is added to the plate in cabinet A, and cDNA in cabinet B. Then, the end prep master mix is prepared and added to the plate in cabinet A. Amplified amplicon pools are then added to the plate in cabinet A. The barcoded amplicons are quantified using the Quantus Fluorometer using the Qubit™ 1X dsDNA HS Assay Kit. The AMII adapter ligation reaction is prepared and then purified by AMPureXP. Station B: the SARS-CoV-2 prepared library is loaded into the MinION for 6 hours. Station C: The PCR is placed in this station, and the plate containing the pool is placed in the thermal cycler for cDNA synthesis and then to run multiplexed PCR. The schematic illustration of the sequencing workflow is shown in Fig 2a.

**Figure 1.**
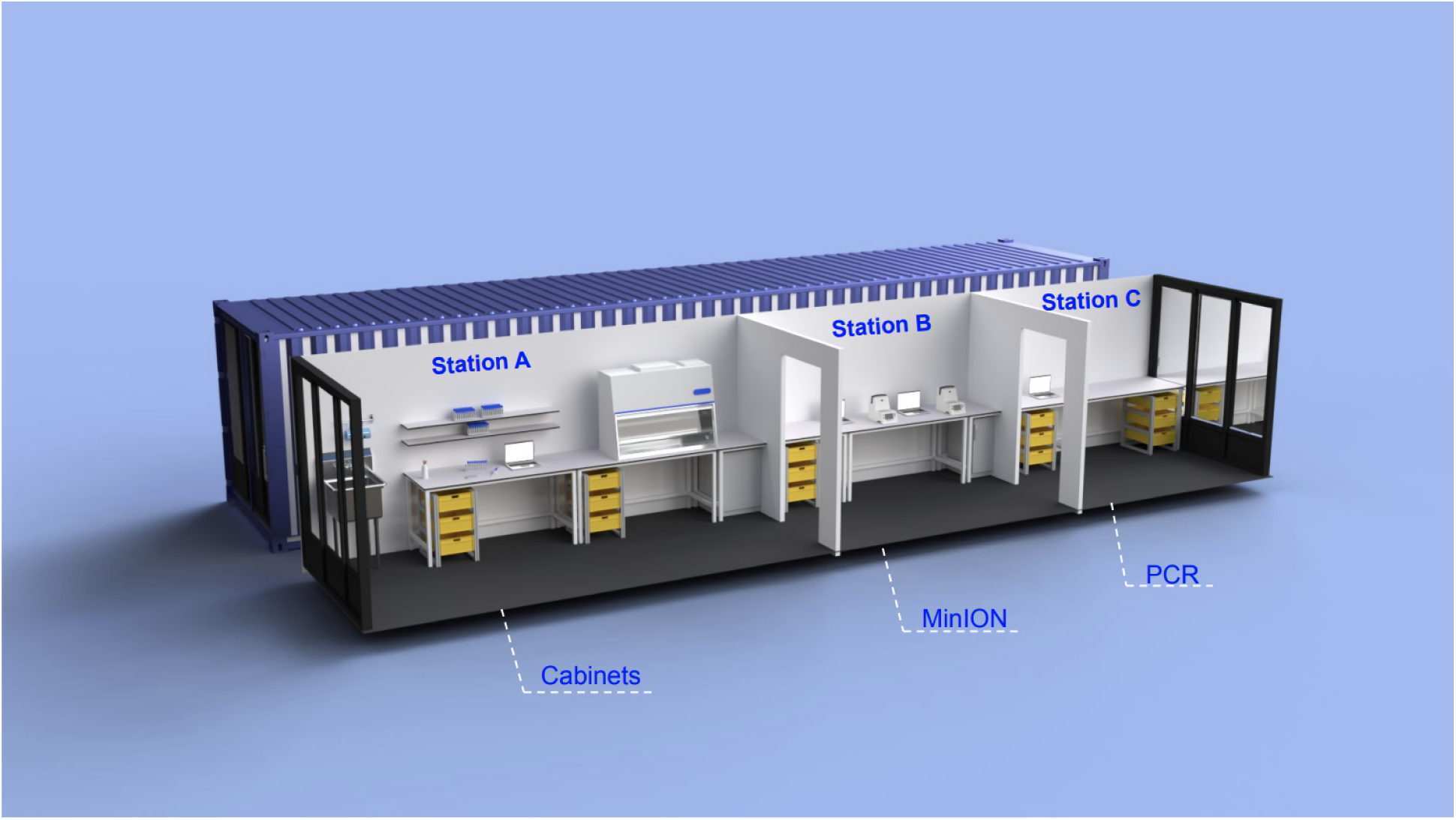
Schematic illustration of sequencing unit. The unit contains three stations. The library preparation is carried out in station A, which then is loaded into the sequencing device in station B. The PCR is in station C.

**Figure 2.**
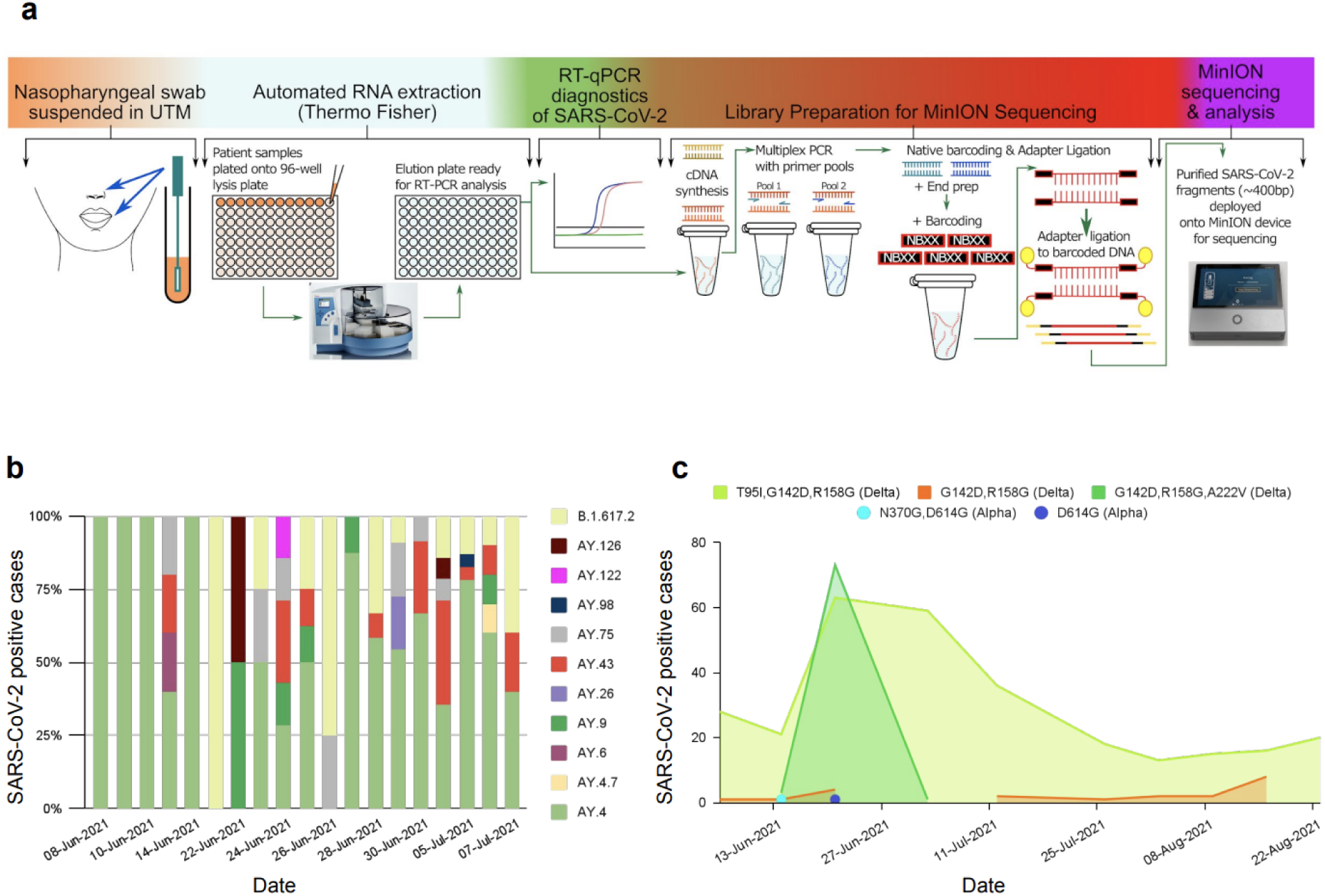
Distribution of lineages in Jersey. **a**.Schematic illustration of workflow for generation of SARS-CoV-2 sequences using ONT. **b**. The lineages of positive cases sequenced are presented in percentage, **c**. The SARS-CoV-2 lineages detected and presented in colors for the period of June 2021 to August 2021.

## Results

### SARS-CoV-2 genomic surveillance in Jersey

In order to identify the SARS-CoV-2 variant introduced into the state of Jersey and to determine the mutation profile, we sequenced the SARS-CoV-2 genome from 154 positive samples from individuals seeking PCR tests between June and July 2021. The sequences were analysed against Wuhan’s reference sequence (NC_045512.2). Genome sequences with >90% genome coverage were analysed. The median Ct value of samples sequenced was 22, and all the sequences identified as the Delta variant lineages B.1.617.2 (18 sequences), AY.4 (92 sequences), AY.4.7 (1 sequence), AY.6 (1 sequence), AY.7 (1 sequence), AY.9 (6 sequences), AY.26 (2 sequences), AY.43 (19 sequences), AY.75 (10 sequences), AY.98 (1 sequence), AY.122 (1 sequence), and AY.126 (2 sequences) (Fig. 2b). The sequences were categorised into two main distinct sublineages based on the mutations. Sub-lineage –I: ORF1a-A1306S, P2046L, P2287S, A2529V, V2930L, T3255I, and T3646A; ORF1b-A1918V; N-G215C, and G18C; S T95I. Sub-lineage -II: ORF1a-P1640L, A3209V, V3718A, and T3750I; ORF1b-T1743N, H1387Y; N-G18C, and G18C; S-A222V, which is known as the Delta sub-clade (21I). Based on mutation analysis, 53% of sequences contained a signature 23,403 A > G (T95I) SNP mutation. Interestingly S: A222V and S: S943T mutations were present in 25% and 9% of sequences, respectively. The mutants with high prevalence (95-100%) in the S gene are T19R, E156-, F157-, R158G, L452R, T478K, D614G, and P681R. Additionally, we noted two mutations of ORF1a that are more prevalent in the Delta plus variant, including nsp6:T181I (ORF1a: T3750I) and nsp4:A446V (ORF1a: A3209V) that were present at frequencies of 25% and 24%, respectively.

In addition to studying the whole genome sequences of 154 samples, we also analysed qualitative data on the mutation profile of 426 cases, which were sequenced in a separate, off-island laboratory (Micropathology Laboratories LTD). The third-party data observed several mutations and identified two variants between June to August 2021. The majority of sequences belonged to the Delta variant (B1.617.2) except for two isolated samples that belonged to the Alpha variant (B.1.1.7). This suggests from June to Sept 2021; the Delta variant became the predominant circulating variant in Jersey. The most prevalent mutations observed in the spike genome were G142D (99.8%), R158G (99.8%), T95I(73.9%), and A222V (18.8%) (Table 1).

**Table 1.** Additional spike mutations were detected in sequences of the Delta genomes.

| Alpha (B.1.1.7) |  | Delta (B.1.617.2) |  |  |  |  |  |
| --- | --- | --- | --- | --- | --- | --- | --- |
| Mutation Prevalence |  | Mutation Prevalence |  | Mutation Prevalence |  | Mutation Prevalence |  |
| D614G | 2 | G142D | 424 | L5F | 2 | K356E | 1 |
| N370G | 1 | R158G | 424 | P174S | 1 | V62L | 1 |
|  |  | T95I | 315 | H655Y | 1 | T22T/I | 1 |
|  |  | K182N | 1 | A27S | 2 | A67A/V | 1 |
|  |  | D138H | 5 | T51I | 1 | D138D/H | 1 |
|  |  | P25T | 1 | G199V | 1 | V615V/A | 1 |
|  |  | A222V | 80 | A522S | 3 | V11F | 1 |
|  |  | D574Y | 1 | D138Y | 3 | S640F | 1 |
|  |  | D138Y | 3 | K558N | 1 | G446V | 2 |
|  |  | A688V | 2 | A706V | 1 | A647S | 1 |
|  |  | R683W/R | 1 | D80Y | 1 | T376I | 1 |
|  |  | D80Y | 1 | K356E | 1 | Y145H | 1 |
|  |  |  |  | T678I | 1 |  |  |

The S: T95I mutant was detected during the summer months, however shortly after the surge of cases, S: A222V was also detected but only for a short period. S: T95I mutant was present during summer, however S: A222V was detected only a short period during the surge in S: T95I cases (Fig. 2c). The surge in S: A222V mutants peaked and dropped rapidly afterwards. Of the 426 cases sequenced, S: G142D and S: R158G mutations were identified as present, except for 2 cases. S: G142D mutation was only present in 2.6% of the cases sequenced in the OpenCell laboratory. The median Ct value of sequences containing S: T95I is 0.5 Ct lower than sequences containing S: A222V.

### Demographics

The data presented in this report covered September 2020 to August 2021, which can be distinguished into two main SARS-CoV-2 outbreaks: the winter outbreak during December 2020 to January 2021 and the summer outbreak during July and August 2021. A total of 598,155 samples were tested for SARS-CoV-2 infection through on-island surveillance screening (community track, trace program, and workforce screening), hospital healthcare, and inbound travel. During both outbreaks, Jersey implemented movement restrictions on residents and mandatory quarantine for inbound arriving passengers. The travel restrictions were lifted in April 2021 after the winter outbreak, and a traffic light region classification was applied for all incoming arrivals. Nonetheless, during the summer months, the number of confirmed positive cases rapidly peaked again. The implementation and lifting of government control measures, dates of major travel and community health interventions are shown in (Fig. 3a). A relatively high incidence of COVID-19 cases was mainly observed in younger age groups. The distribution of cases based on gender illustrated that more males than females received positive SARS-CoV-2 results (50.9% vs 49.1%) from August 2020 to August 2021. The infection rate in females was slightly higher (4.3%) than males during the winter outbreak. However, this was slightly changed during the summer outbreak by increasing males (2.9%) diagnosed with COVID-19 (Fig. 3b).

**Figure 3.**
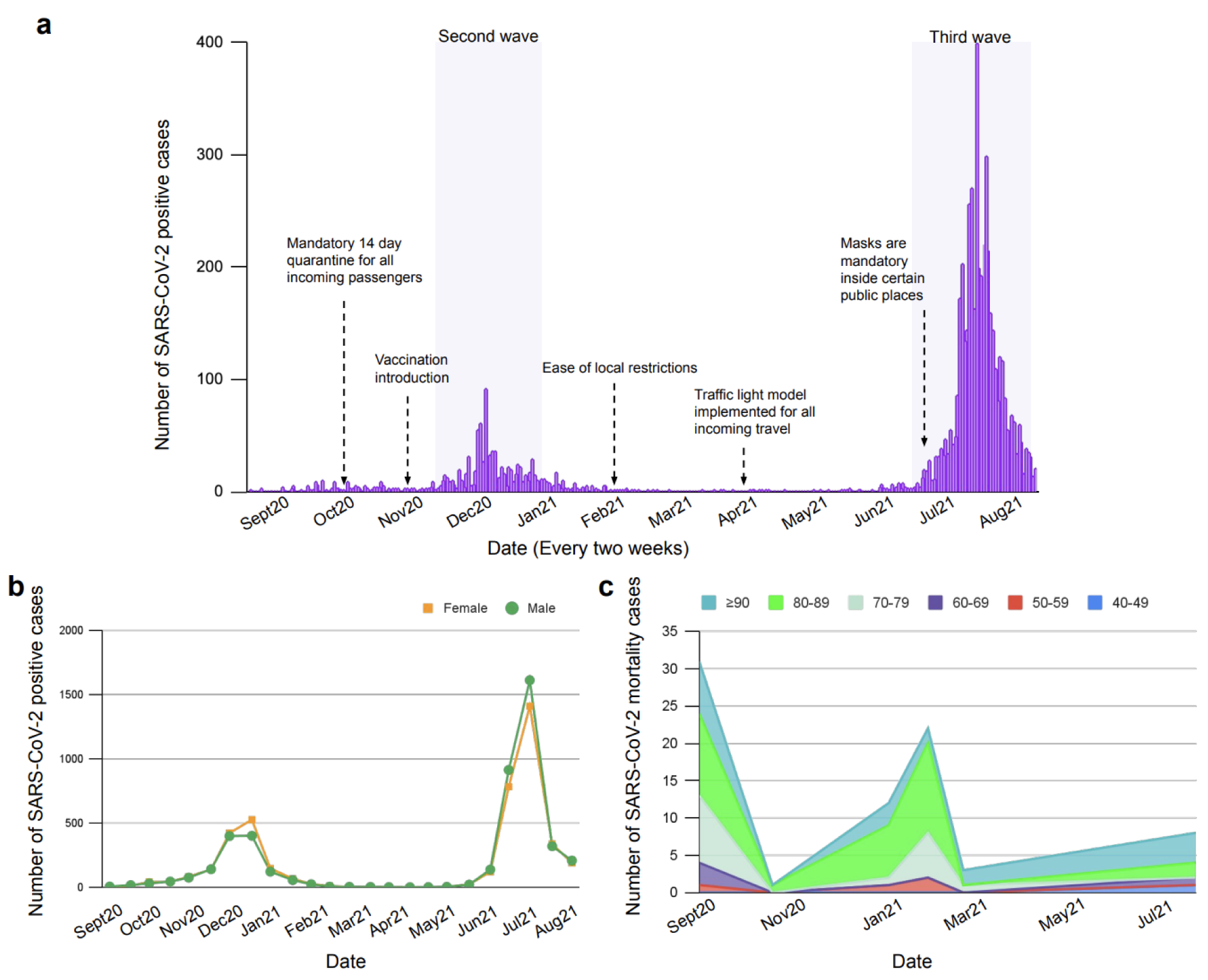
The positive and mortality cases of SARS-CoV-2 in Jersey identified during September 2020 and August 2021. **a**. SARS-CoV-2 winter outbreak of 2020 took place during Dec-Jan 2021 and the summer outbreak during July and Aug 2021. The implementation and lifting off government control measures is shown in the graph. The dates of major travel and community health interventions are indicated with arrows, **b**. Proportion of SARS-CoV-2 tests by sex during September 2020 to August 2021. **c**. The distribution of mortality cases by age during October 2020 to August 2021..

### Association of age and sex with COVID-19 mortality

Epidemiological factors investigated for COVID-19 mortality were sex and age. In Jersey, 77 deaths were registered during the pandemic, with 55 confirmed deaths resulting from SARS-CoV-2. An age-dependent mortality rate had been demonstrated, with no death recorded among those younger than 39. The risk of mortality increases incrementally, affecting the 40–49 years old age group (1.3%), among those aged 50–59 years (5.5%), at age 60–69 years (5.2%), age 70-79 (22.1%), and with the highest mortality rate in those age ≥80 years (66.3%) (Fig. 3c). The median age of deceased patients related to SARS-CoV-2 was 78 years. Additionally, the mortality rate was 21% higher in males than females.

### Distribution of positive cases of SARS-CoV-2

During September 2020 and August 2021, 598,155 tests identified 8,950 (1.5%) SARS-CoV-2 positive cases. Overall, 283,357 (47.4%) tests were administered as part of on island surveillance screening, 21,168 (3.5%) tests in individuals with symptoms at the time of testing, and 293,630 (49.1%) tests for inbound travellers. The distribution of positive cases with or without a symptom from September 2020 to August 2021 is illustrated in (Fig. 3a). Most individuals hospitalised were residents of care homes (Fig. 3b). During the winter outbreak, 200 hospitalisations were recorded due to SARS-CoV-2 infection, of which 33 had fatal outcomes; this was reduced to 50 hospitalisations and 8 deaths during the summer outbreak (Fig. 3b). The number of positive cases from inbound travel also increased during the summer holiday compared to winter. The health care workers who tested positive during the winter outbreak counted 157 (20.9%) of total positive cases and later reduced to 135 (6%) during the summer outbreak. Symptomatic cases accounted for 32.6% of positive cases from January to July 2021. The mean age of SARS-CoV-2 symptomatic cases was 33 years old (interquartile range (IQR), 20-45). The 995 (45.4%) of total positive cases were individuals with symptoms during the summer outbreak, compared to 143 (19%) in the winter outbreak (Fig. 3c). The self-reported symptoms by 117 individuals who tested positive were analysed to assess the prevalence. The most frequent symptoms experienced at the time of the test were headache (55%), sore throat (19%), runny/stuffy nose (17%), fatigue (14%), and persistent cough (14%).

### Effect of vaccination and age by viral load

Between January 2021 and September 2021, the viral load of 6540 SARS-CoV-2-infected individuals was investigated. All Ct values were obtained from RT-qPCR diagnostic assays conducted on samples at the OpenCell laboratory. The viral loads of cases infected during January 2021 (n=307, Ct=25.3 for the S gene, IQR, 20.97-31.18) were lower than individuals infected during July 2021 (n=4668, Ct=23.79 for the S gene, IQR, 20.57-29.91). The median age of the individuals who tested positive was 36 years. The age distribution of confirmed positive cases at the end of December 2020 is opposite to July during the summer outbreak. The positive cases shifted considerably towards older age groups during the winter outbreak. However, during July 2021, the confirmed cases among the elderly continuously decreased (Fig. 3d). To determine the effect of age on the viral load, 822 Ct values from positive samples for both target genes arranged by different classes of age (>10, 10-19, 20–29, 30–39, 40–49, 50–59, 60–69, 70–79, and ≥80), were analysed and were found to be statistically significant (S gene p= 0.02, E gene p=0.01). Mean Ct value was also noticeably lower by 0.6 Ct for E gene and 1 Ct for S gene in the age group ≥80 (Table. 2). The mean Ct of the 70-79 age group was 1.9 and 1.8 Ct lower than the mean of all the E and S genes, respectively. The mean Ct of age group 60-69 was higher by 1.7 for the S gene and 2.9 for the E gene compared to mean Cts of all specimens, suggesting a lower viral load in the samples of this age category. Almost half of the positive cases (49.1%) were observed in the younger age group (10-29). A higher viral load was observed among individuals over 70 years of age. The lowest Ct value was 13 for seven samples and the highest 43 (Supplementary Fig S4). The vaccination status of these individuals is unknown. By March 2021, the Jersey Government announced that 100% of all ≥80s had been fully vaccinated against SARS-CoV-2. At the start of July 2021 (summer outbreak), the same body reported that 51% of the population had both doses of a SARS-CoV-2 vaccine administered.

**Table 2.** Mean, and median Ct values for 822 specimens were reported as positive for SARS-CoV-2 for target E and S genes.

| Age group | No. of Specimens | Target 1 (S-Gene) |  |  |  | Target 2 (E-Gene) |  |  |  |
| --- | --- | --- | --- | --- | --- | --- | --- | --- | --- |
|  |  | Mean CT | Median CT | IQR | P Value | Mean CT | Median CT | IQR | P Value |
| All specimens | 822 | 25.6 | 25.1 | 21.3 - 30.0 | 0.01 | 26.8 | 26.5 | 21.2 - 30.8 | 0.02 |
| <10 | 29 | 26.0 | 25.9 | 21.6 - 29.2 |  | 26.6 | 26.5 | 21.9 - 29.6 |  |
| 10-19 | 193 | 25.8 | 25.8 | 21.1 - 30.4 |  | 26.7 | 26.5 | 22.0 - 31.5 |  |
| 20-29 | 211 | 25.8 | 25.1 | 21.2 - 30.0 |  | 26.8 | 26.1 | 22.2 - 31.3 |  |
| 30-39 | 104 | 25.7 | 25.0 | 20.9 - 31.1 |  | 27.0 | 26.5 | 22.1 - 32.4 |  |
| 40-49 | 108 | 24.9 | 24.6 | 20.7 - 29.9 |  | 26.5 | 25.2 | 21.5 - 30.8 |  |
| 50-59 | 100 | 26.3 | 25.4 | 22.5 - 30.9 |  | 27.4 | 27.0 | 23.1 - 32.7 |  |
| 60-69 | 35 | 27.3 | 28.4 | 22.5 - 32.6 |  | 29.1 | 29.4 | 23.9 - 34.7 |  |
| 70-79 | 19 | 23.8 | 20.9 | 19.6 - 28.5 |  | 24.9 | 23.5 | 20.3 - 30.1 |  |
| ≥80 | 23 | 24.6 | 24.4 | 21.3 - 27.7 |  | 26.2 | 25.5 | 13.9 - 24.5 |  |

We next investigated the effect of vaccination (1 or 2 doses) on viral load reduction by comparing it to unvaccinated individuals in the same age group who arrived in Jersey (n=796) between December 2020 to August 2021. 1,129 (0.33%) of 283,357 inbound passengers confirmed positive upon arrival (day 0), 73 confirmed positive on day 5 test, and 93 were positive on day 8 or day 10 tests. 29.7% of infected passengers were vaccinated, mostly older individuals, as expected from the age-prioritisation vaccine roll-out in the UK. The individuals aged ≥40 had a higher viral load among infected unvaccinated groups than vaccinated, except age groups 50-59, which showed a higher viral load among vaccinated (Table. 3). The 30-39 had a slight effect between vaccinated and unvaccinated groups. However, stratifying Ct values based only on vaccination status showed slight differences, with lower viral load in the vaccinated group (Ct=30.47, Ct=30.02, respectively).

**Table 3.**
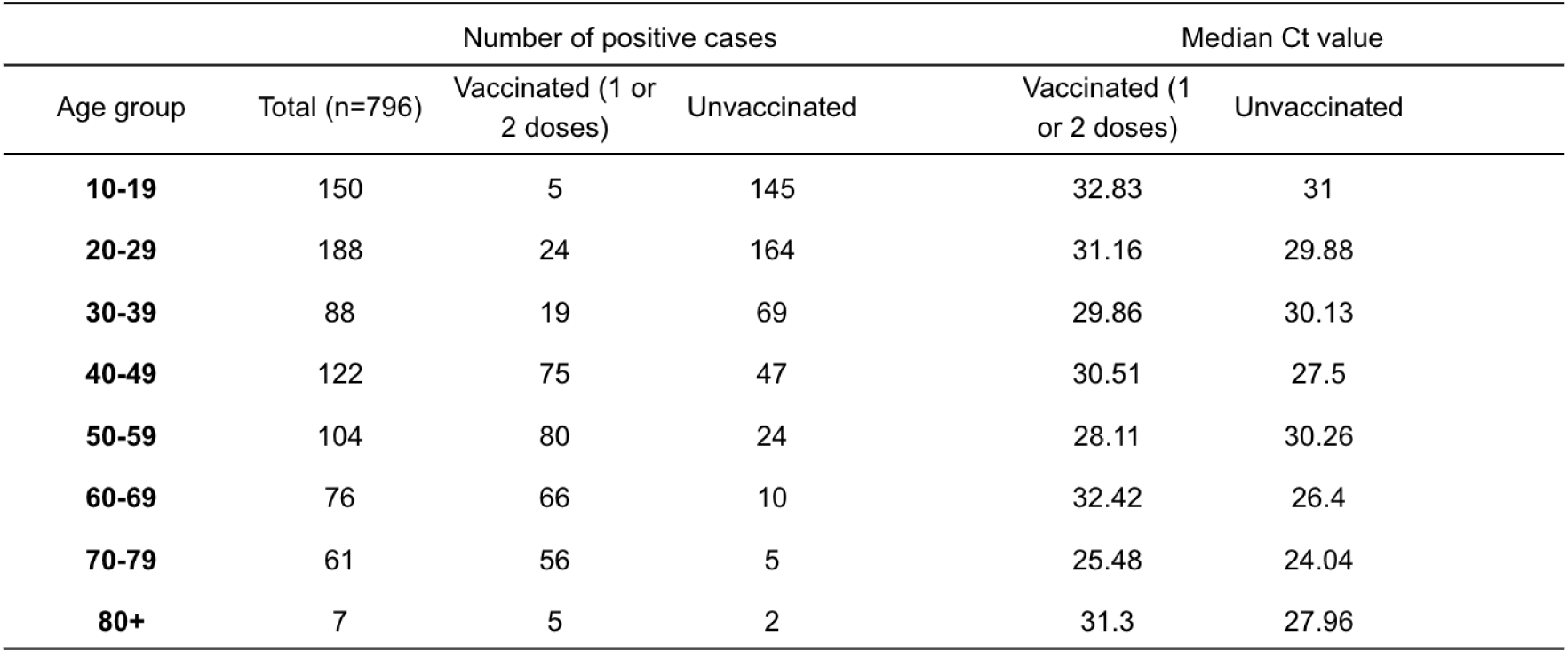
Viral loads of vaccinated and unvaccinated positive SARS-CoV-2 individuals. The most significant effect was observed among the 60-69 age group, with the unvaccinated showing higher viral load.

### Direct contact test and trace

A contact tracing strategy was carried out to identify people in direct contact with individuals who tested positive during June and July 2021. 11,220 individuals were tested as part of the contact tracing strategy on days 0, 5, and 10. 911 (8.1%) tested positive in test 1 (day 0), 591 tested positive on the second test (day 5), and 185 individuals continued to test positive on test 3 (day 10). The viral load of direct contacts was assessed by age and date, displayed in Fig. 5. The age of infected individuals was higher during the first test. The individuals who remained positive on the second and third tests were younger than the median age in the first test on day 0. The viral load had a positive, strong significant correlation with the age of the direct contact positive cases (r = 0.82, P < 0.04).

**Figure 4.**
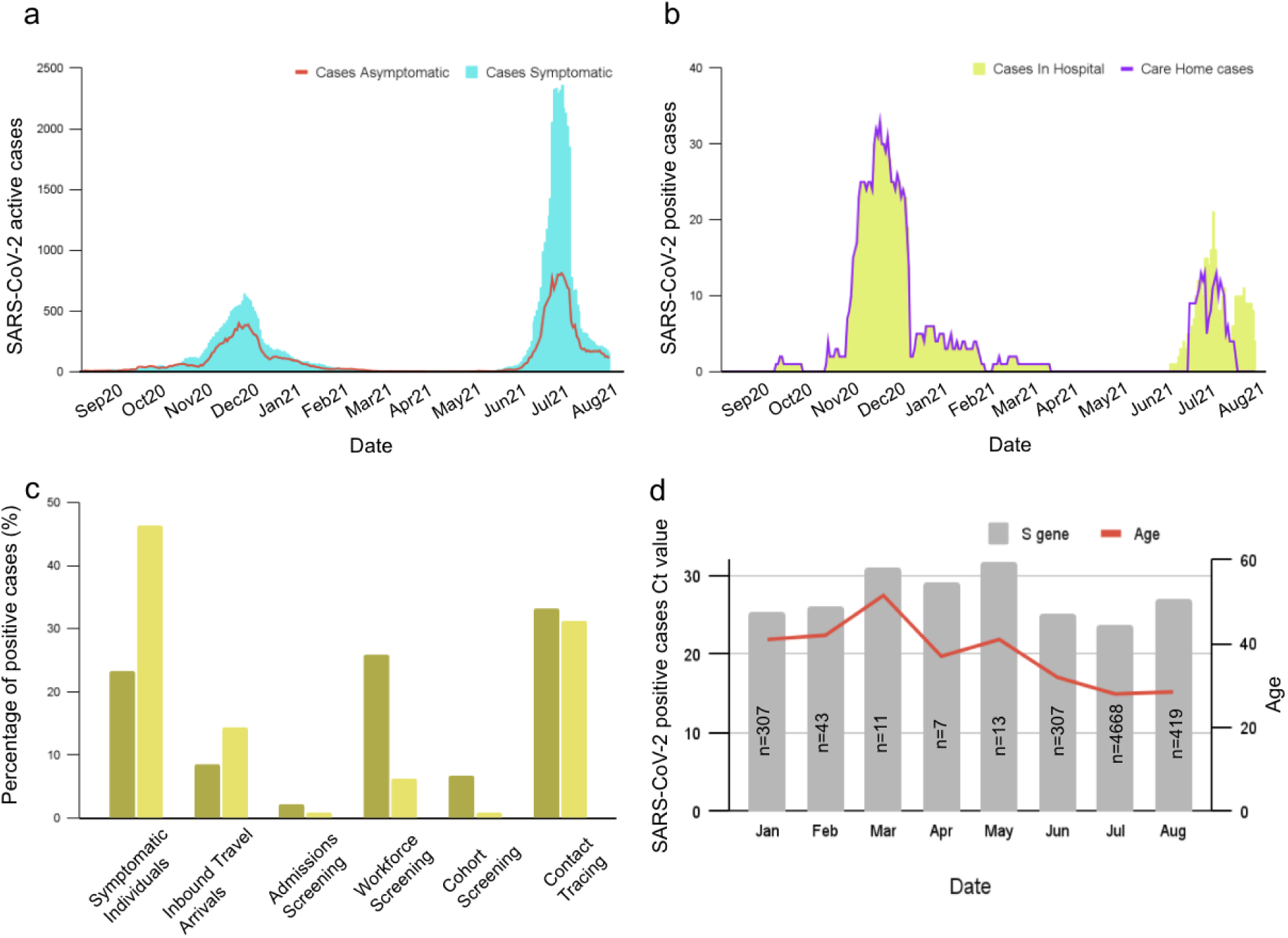
The SARS-CoV-2 positive cases distribution based on reason to seek test. **a**. The distribution of SARS-CoV-2 active cases based on presence or absence of symptoms between Sep 2020 to Aug 2021. **b**. The number of SARS-CoV-2 positive cases in hospitals and care homes, **c**. Proportion of SARS-CoV-2 tests by platform utilized, **d**. Ct values for the S and E genes (n=6540) together with age of positive individuals were presented during January 2021 to September 2021.

**Figure 5.**
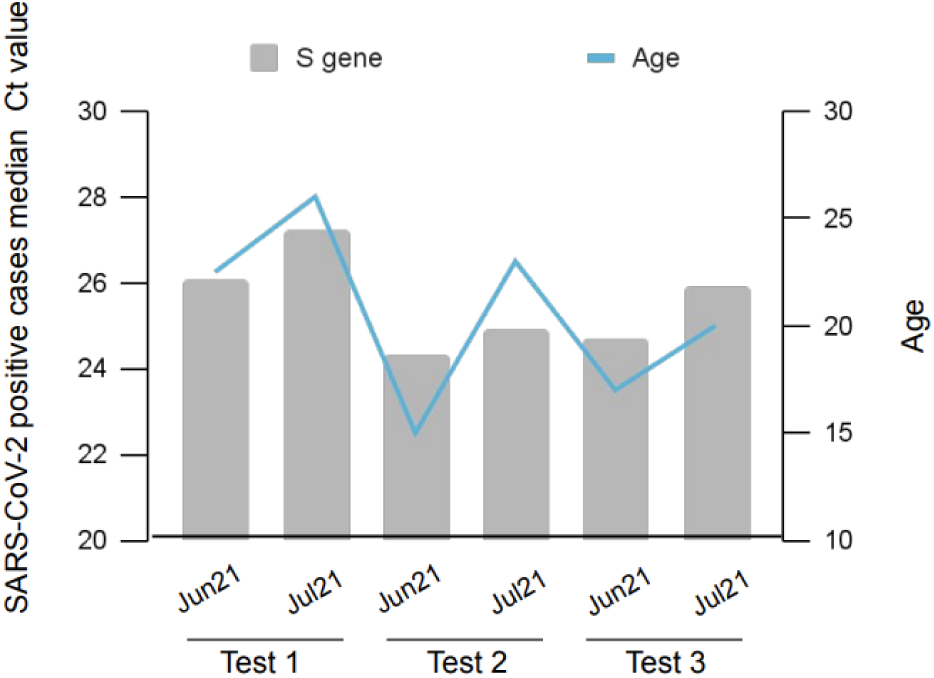
The contact tracking strategy for June.and July 2021. Three tests over 10 days was crried out to identify people who have been in direct contact with indivuals. The age and Ct values of infected indivuals are illustrated in the graph.

## Discussion

We studied rapid SARS-CoV-2 genome sequencing to assess amino acid changes in the virus and focused on the epidemiology in a well-controlled and monitored setting on the island of Jersey. The ONT device was used for sequencing, which offered a portable and cost-effective solution. The turnaround time of sequencing using the ONT devices is straightforward and fast. The library preparation takes about 12 hours and 6 hours to run the library on the device to receive the fast5 or fastq files. The rapid sequencing of SARS-CoV-2 enables fast identification of new variants in the local laboratories rather than days in overseas laboratories.

This study aimed to investigate the mutation profile of positive individuals in Jersey during the summer outbreak. The genome sequencing carried out in the OpenCell laboratories showed the Delta variant sub-clade (21I and 21J) as the dominant variant circulated in the community during the summer outbreak. Two main sublineages were identified based on the signature mutations in the spike (S: A222V and S: T95I). The prevalence of S: A222V mutant declined rapidly and presented only briefly (12th June to 7th July 2021). S: A222V was first identified and expanded during early summer 2020 in Spain and was characteristic of the B.1.177 lineage ^33^. S: T95I was found in more than half of the sequences during summer 2021. Interestingly, this mutation is also associated with the Omicron variant. The viral load of samples containing T95I are slightly higher than the samples containing A222V. The increased viral load of this sublineage is associated with the presence of NTD mutation S: G142D. S: G142D mutation was present in 2.6% of our sequenced samples; however, it was observed in all the Delta cases identified in the qualitative data. The variation in the G142D mutation in the Delta genome is caused by the dropout of a sequencing amplicon ^34^. The reappearing of the mutations, including S: A222V and S: T95I in new emerging variants, may be attributed to the genetic background of the population where the mutation originated. Furthermore, the impact of population genetic processes such as founder effects that alter genetic variations could be a significant factor in the presence of some mutations in several lineages. S: L452R (96.5%) and S: T478K (100%) were almost always present in all genome sequences of the sublineage I and II list of mutations, which is known to cause structural changes that increase spike stability and disrupt the RBD interaction with the ACE2 receptor, resulting in a more infectious and high-risk SARS-CoV-2 ^35^. The distribution of infected cases sharply increased from December 2020 to January 2021 and later declined slowly. By March 2021, the Jersey Government announced all ≥80s had been fully vaccinated, and at the start of July 2021, 51% of the population had both doses of a SARS-CoV-2 vaccine administered. Enhancing the vaccination drive and immunising populations quickly showed to be the most effective strategy for preventing future deadly waves. The viral load of the vaccinated group showed to be lower than that of unvaccinated individuals, which may imply that vaccination affects the decline in the viral load faster and reduces infectiousness. However, the Delta variant was the dominant variant worldwide during summer 2021, which has higher transmissibility than the Alpha variant and led to a surge in infected cases ^36^. The outbreak during the summer corresponded to a relatively young age group with rapid growth in daily confirmed incidence of SARS-CoV-2 infection. As expected from the age prioritisation of the vaccine program, more younger individuals were unvaccinated during the Delta variant circulation (summer 2021). The escalation of inbound travel, the peak holiday season and young individuals moving more during summer holidays may have partly facilitated the rapid transmission of the Delta variant. Thus, leading to the surge in the number of observed infected cases compared to the winter outbreak. During this period, the number of hospitalisations and deaths has substantially dropped compared to winter, implying that population vaccination reduces the number of hospitalisations and mortality associated with SARS-CoV-2 ^37–39^. The study of positive cases in Jersey illustrated that A slightly higher percentage of infected cases was observed in females during the winter outbreak with no statistical significance. However, the overall proportion of SARS-CoV-2 infected cases is higher in males than females, which may suggest the SARS-CoV-2 virus has sex-specific differences in infectivity. The slightly higher positive cases in females could partially be because they are more likely to follow the guidelines during the pandemic and, therefore, more likely to participate in regular screening tests ^40^. The mortality rate was 1.5-fold higher in males, suggesting male sex is a risk factor associated with SARS-CoV-2 mortality. The gender gap observed may point to an underlying biological mechanism. Biological sex differences affect immune responses that lead to differential susceptibility and disease progression mechanisms and distinct responses to the vaccine ^41,42^. Males have been recorded to have more significant expression of ACE2 receptors ^43,44^, impacted by X chromosome and sex-related hormonal levels ^45^. Moreover, social-cultural constructs and behavioural differences may also play fundamental roles in mediating the development of severe COVID-19 outcomes in males ^46,47^. In this study, the mortality rate in patients older than ≥80 years was 66.3% of total cases, indicating age is a strong predictor of mortality for SARS-CoV-2. Aged immune systems combined with other age-related complications, such as reduced reserve capacity of vital organs and comorbidities, all of which make older adults susceptible to a more severe disease course or death when responding to COVID-19 infection ^48–52^. All population ages are prone to COVID-19 infection, with the average age of infection being 36 years in Jersey. This study examined associations between SARS-CoV-2 viral load and the age of confirmed COVID-19 cases across seven age groups. Higher viral loads were observed in adults aged ≥70, suggesting that viral RNA levels are higher in older age groups than for younger positive individuals. This observation is further supported by several other studies that found a higher viral load among the elderly groups ^51,53,54^. The viral load was higher during Delta prevalence in the summer outbreak, which may partially explain this variant’s rapid and intense transmission ^21,55,56^. The contact trace investigations over 10 days showed that younger individuals stayed positive for longer. One reason could be that nearly all older adults were vaccinated during summer 2021, indicating that vaccination affects the faster viral decline.

This study has several limitations. The samples selected for sequencing only contained Ct values below 30, as it is difficult to sequence very low viral load samples. The sex of individual positive cases was missing, and only total positive cases for each month were considered. The vaccine status of positive cases in the community was unknown and therefore was not considered in the Ct value study. Infected inbound passenger individuals with either 1 or 2 doses of vaccine were categorised in one group. We compared the median Ct value of the summer outbreak to winter; however, the community’s variants during summer can not be confirmed. While the dominant variant present during winter in the UK is the Alpha variant, this can not be confirmed as no sequencing has been carried out on samples during that time.

## Conclusions and Future Work

Future work is required to focus on a more longitudinal approach to study and compare the Delta variant viral load to other variants. Further studies are required to improve the limitations of the ONT, such as detecting InDel, which remains a problem. The base-calling accuracy requires better performance and shorter turnaround time, which implies improved algorithms.

In conclusion, we present data suggesting age and male sex are risk factors for mortality from COVID-19 infection. The infected older age (70+) individuals had a higher viral load. The positive summer cases were among younger individuals, with symptomatic cases escalating during the summer outbreak compared to winter. However, the number of care home cases, hospitalisations and deaths were substantially decreased during summer. 30% of the positive cases among travellers were vaccinated and showed a lower viral load across the spectrum of age. Finally, the younger infected individuals stayed positive for longer, and the age of SARS-CoV-2 infected individuals who tested positive was reduced from winter to summer.

## Data Availability

All data produced in the present study are available upon reasonable request to the authors.

## Author contributions

SFB conceived and designed the analysis and carried out the genome sequencing, quality control, and genome assembly. AI polished and checked the quality of genome sequences. AT categorised and classified Ct value data. SFB and DJ wrote the manuscript. TM and HS supervised the project from conducting the research to writing the manuscript. All authors critically corrected and revised the manuscript.

## Acknowledgements

We thank G. Kerr, K. Osborne. A. Hoogkamer, J. Andron, B. Edwards, and V. Morel for their invaluable help in the transfer of the government of Jersey SARS-CoV-2 data, which was central to the achievement of this work, J. Perez, E. Bransden, and members of COVID-19 diagnostic laboratories of OpenCell for their help and excellent support throughout the sequencing work. We thank Stanhope Plc for facilitating and accommodating shipping container laboratories on their premises.

## Supplementary Materials

See supplementary materials with figures S1-4.

## Notes

### Competing Interest Statement

The authors have declared no competing interest.

### Funding Statement

This study was funded by Open Cell.

### Author Declarations

Ethics Committee of Jersey General Hospital (Bailiwick of Jersey) gave ethical approval for this work.

