## Supplementary figures 1-4 for "Rapid genome surveillance of SARS-CoV-2 and study of risk factors using shipping container laboratories and portable DNA sequencing technology"

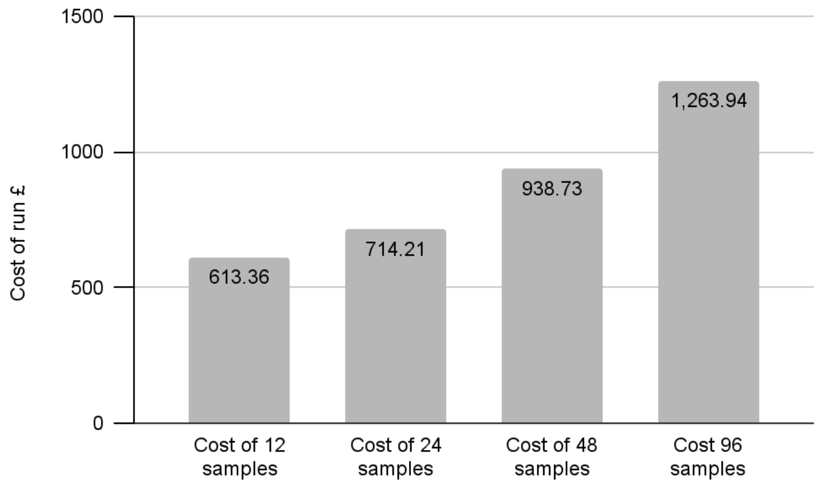


**Figure S1. The cost of performing a sequencing run has been compared between runs with different numbers of samples for sequencing using the nanopore Minion.** The x-axis represents the number of samples in a run, and the y axis the cost in pounds. The costs are calculated based on using the flow cell three times by washes after each use. The cost of one sample is more expensive, and it is cost-effective when more numbers of samples are analysed.

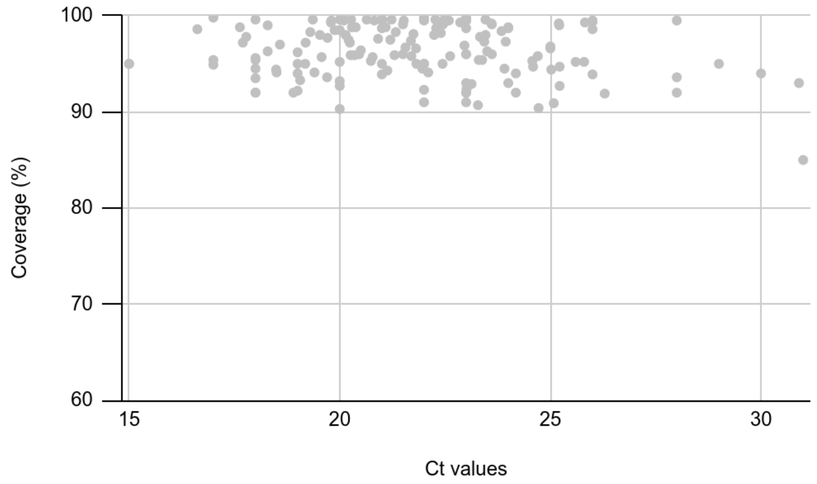


**Figure S2. SARS-CoV-2 genome coverage for different Ct values**. Sequence coverage of 167 SARS-CoV-2 positive cases sequenced is illustrated with the corresponding Ct number. The lower the viral load (i.e., the higher the Ct value), the lower the sequence coverage obtained. It is difficult to obtain sufficient coverage on the samples with Ct numbers above 30.

| Regions (ORF) | Amino acid mutations Sub-lineage I | | | Amino acid mutations Sub-lineage II | | |
|---|---|---|---|---|---|---|
| | Site | No. | Type | Site | No. | Type |
| 1a | 1306 | 115 | A→S | 1640 | 43 | P→L |
| | 2046 | 128 | P→L | 3209 | 41 | A→V |
| | 2287 | 124 | P→S | 3718 | 42 | V→A |
| | 2529 | 102 | A→V | 3750 | 43 | T→I |
| | 2930 | 128 | V→L | | | |
| | 3255 | 126 | T→I | | | |
| | 3646 | 128 | T→A | | | |
| 1b | 1918 | 128 | A→V | 1743 | 32 | T→N |
| | | | | 1387 | 36 | H→Y |
| S | 95 | 90 | T→I | 222 | 43 | A→V |
| N | 215 | 128 | G→C | 18 | 36 | G→C |

**Figure S3. Characterisation of two sub-lineages observed in the Delta SARS-CoV-2 variant (n=167).** The amino acid site, prevalence and type of mutations observed in different genes are illustrated in the table above.

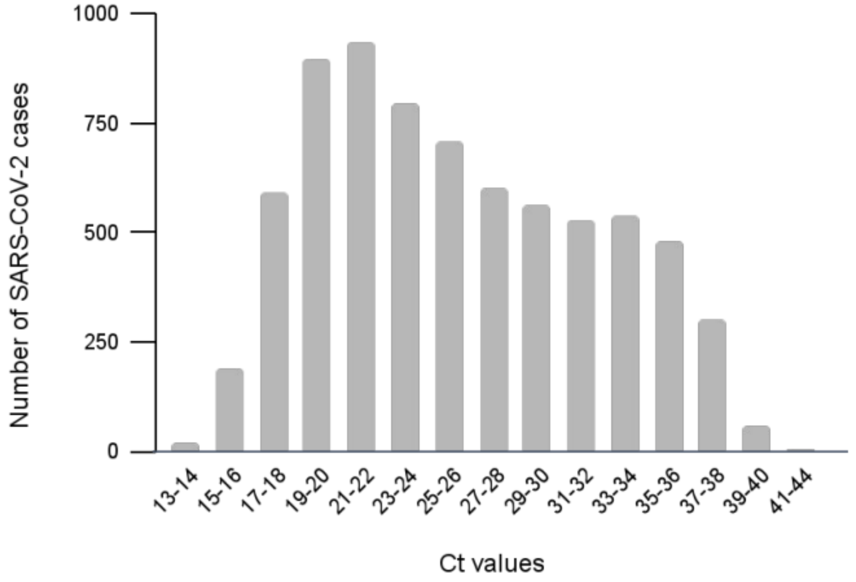


**Figure S4. The distribution of Ct values of the S gene.** The Ct values of samples from December 2020 to September 2021.
